# RNA polymerase inhibitor enisamium for treatment of moderate COVID-19 patients: a randomized, placebo-controlled, multicenter, double-blind phase 3 clinical trial

**DOI:** 10.1101/2022.08.21.22279036

**Authors:** O. Holubovska, P. Babich, A. Mironenko, J. Milde, Y. Lebed, H. Stammer, L. Mueller, A.J.W. te Velthuis, V. Margitich, A. Goy

**Author notes:** contributed equally (O. Holubovska, P. Babich).

## Abstract

Enisamium (trade name Amizon^®^ MAX) is an orally available therapeutic that inhibits influenza A virus and SARS-CoV-2 replication *in vitro* and improves influenza patient recovery. We evaluated the clinical efficacy of enisamium treatment combined with standard care compared to standard care plus treatment with a placebo control in adult, hospitalized patients with moderate COVID-19 requiring external oxygen. Hospitalized patients with laboratory-confirmed SARS-CoV-2 infection were randomly assigned to receive either enisamium (500 mg per dose, 4 times a day) or a placebo. All patients received standard of care as deemed necessary by the investigator and the health status of each patient. The primary outcome was an improvement of at least two points on an 8-point, modified WHO severity rating (SR) scale within 29 days of randomization. A total of 592 patients were enrolled and randomized between May 2020 and March 2021. Patients with a baseline SR of 4 were divided into two groups: 142 (49.8%) were assigned to the enisamium group and 143 (50.2%) to the placebo group. No differences were observed between the safety or patient tolerability profiles of the enisamium and placebo treatment. Analysis of the population showed that if patients were treated within 4 days of the onset of COVID-19 symptoms (n = 33), the median time to improvement was 8 days for the enisamium group and 13 days for the placebo group (p = 0.0051). For patients treated within 10 days of the onset of COVID-19 symptoms (n = 154), the median time to improvement was 10 days for the enisamium group and 12 days for the placebo group (p = 0.002). Comparison of groups using a stratified one-sided Logrank criterion (adjustment using stratification by age categories: “<40 years”, “40-<65 years” and “≥65 years”) showed statistically significant differences between the groups (p = 0.00945, one-sided). Our findings suggest that enisamium is safe to use in COVID-19 patients, and that enisamium treatment offers a clinical benefit when given to patients with moderate COVID-19 requiring supplementary oxygen, if enisamium is given within 10 days of the onset of symptoms. This trial was registered with ClinicalTrials.gov under NCT04682873.

## INTRODUCTION

Severe acute respiratory syndrome coronavirus 2 (SARS-CoV-2) was identified in December 2019 as the cause of a respiratory illness designated coronavirus disease 2019 (COVID-19).^1^ Various new and repurposed antiviral drugs have been considered as treatment for COVID-19 and evaluated in clinical trials, including remdesivir, favipiravir, azithromycin, hydroxychloroquine, and ritonavir among others.^2, 3^ To date, only the antivirals remdesivir (Veklury), a combination of nirmatrelvir with ritonavir (Paxlovid), and molnupiraivir (the prodrug of beta-D-N4-hydroxycytidine) have received emergency use approval (EUA) for COVID-19 treatment by the FDA and EMA and few repurposed drugs have shown clinical efficacy in reducing mortality, the need for mechanical ventilation or improving the clinical status of COVID-19 patients admitted to a hospital.^4–6^

Early in the COVID-19 pandemic, the World Health Organization (WHO) identified enisamium (4-(benzylcarbamoyl)-1-methylpyridinium, trade name Amizon^®^ MAX) as a candidate drug for the treatment of COVID-19. Enisamium is licensed for the treatment of influenza in 11 countries, and for the treatment of COVID-19 (in hospitalized patients with moderate disease) in Ukraine.^7^ A recent study found that enisamium is efficiently hydroxylated in humans and human lung cells to an active compound, called VR17-04, that inhibits the activity of the influenza virus RNA polymerase.^8^ A phase 3 clinical trial provided evidence that enisamium treatment reduces viral shedding and improves patient recovery in influenza patients.^8^ Toxicology studies found no genotoxic effects of enisamium in an Ames test, no clastogenic activity in human peripheral lymphocytes with and without metabolic activation, no effect on the incidence of chromosome aberrations at any concentration, and no clinical signs of toxicity or cytotoxicity in bone marrow or micronuclei in Wistar rats at any dose. In addition to these reports, a recent *in vitro* study demonstrated that enisamium can also inhibit SARS-CoV-2 replication in cell culture, while molecular dynamics simulation suggested that the active compound can bind to the catalytic site of the SARS-CoV-2 RNA polymerase non-structural protein 12.^9, 10^

Here, we report the evaluation of the efficacy and safety of enisamium in patients hospitalized with COVID-19. We describe a phase 3 clinical trial that was performed in 14 clinical centers and the data which led to the approval to use enisamium for the treatment of COVID-19 (in hospitalized patients with moderate disease) in Ukraine and several other countries. The data reported here indicate that standard care in combination with enisamium treatment outweighs the effectiveness of standard care in combination with placebo treatment in patients with moderate COVID-19 requiring additional oxygen.

## METHODS

### Study design

This multicenter, double-blind, placebo-controlled, randomized, comparative, parallel study was conducted from 15 May 2020 to 26 March 2021 in 14 clinical centers across Ukraine. The protocol and materials of the clinical trial were approved by the CEB (Ministry of Health [MOH] of Ukraine; approval number 2949, December 18, 2020) and the Ethics Commissions at the treatment and prevention facilities where the study was conducted. The study was conducted in accordance with the Declaration of Helsinki, the International Principles for Clinical Trials (ICH GCP), the current legislation of Ukraine, and the approved study protocol. All patients gave written informed consent to participate in the study.

### Criteria for inclusion or exclusion

A total of 592 patients were assessed based on the Modified WHO Ordinal Scale for Clinical Status Patient state (Table 1). For inclusion in the study, each patient had to meet the inclusion criteria listed in Table 2. Patients were excluded from participation in the study if they met any of the criteria listed in Table 3.

**Table 1.**
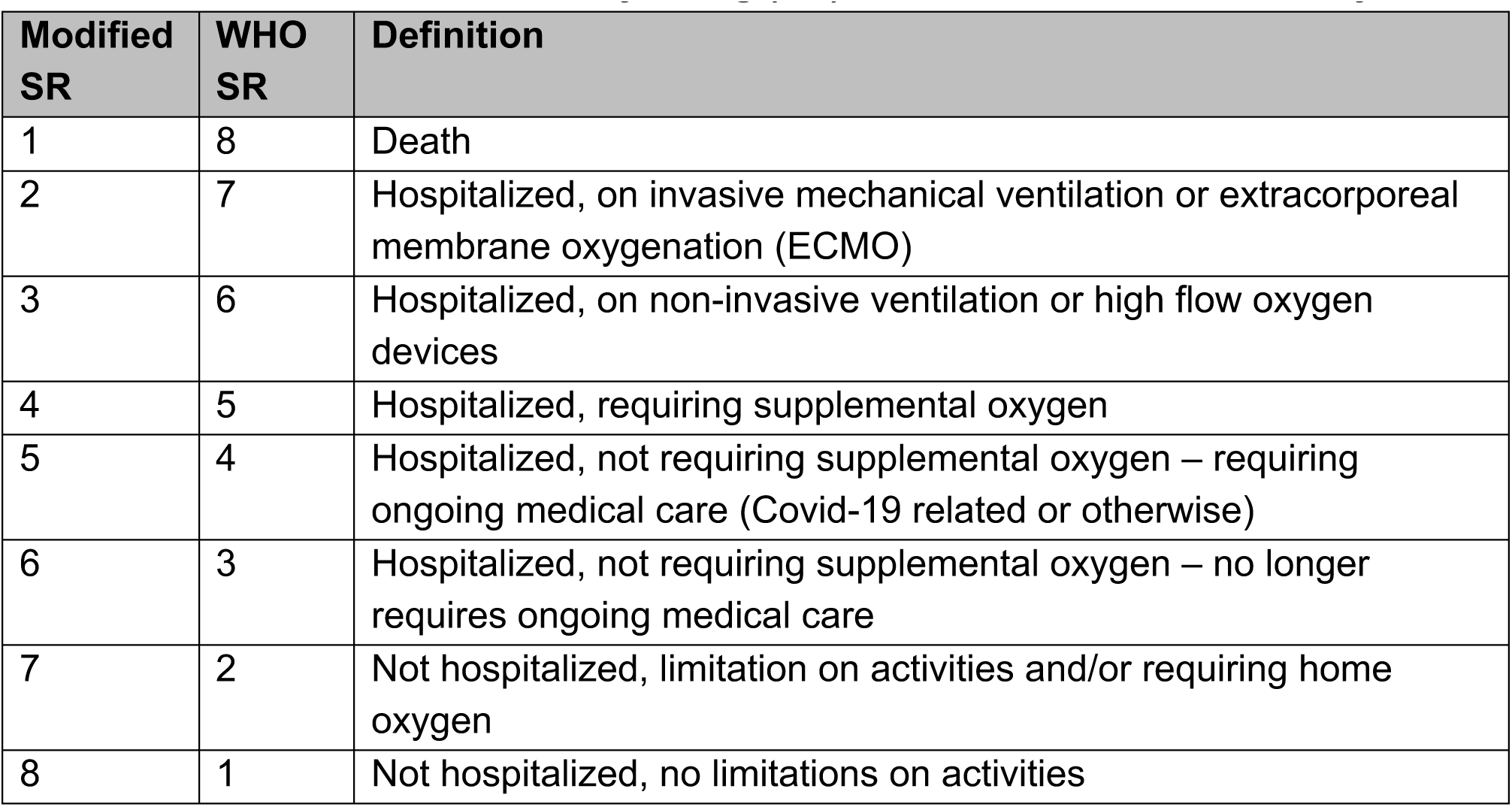
The modified WHO severity rating (SR) scale was used for this study:

| Modified SR | WHO SR | Definition |
| --- | --- | --- |
| 1 | 8 | Death |
| 2 | 7 | Hospitalized, on invasive mechanical ventilation or extracorporeal membrane oxygenation (ECMO) |
| 3 | 6 | Hospitalized, on non-invasive ventilation or high flow oxygen devices |
| 4 | 5 | Hospitalized, requiring supplemental oxygen |
| 5 | 4 | Hospitalized, not requiring supplemental oxygen – requiring ongoing medical care (Covid-19 related or otherwise) |
| 6 | 3 | Hospitalized, not requiring supplemental oxygen – no longer requires ongoing medical care |
| 7 | 2 | Not hospitalized, limitation on activities and/or requiring home oxygen |
| 8 | 1 | Not hospitalized, no limitations on activities |

**Table 2:**
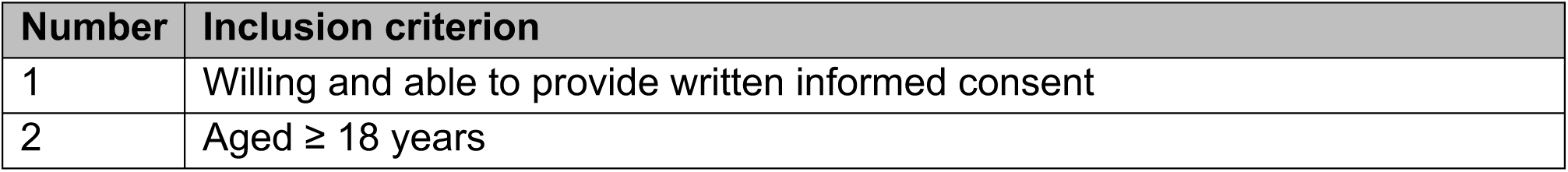
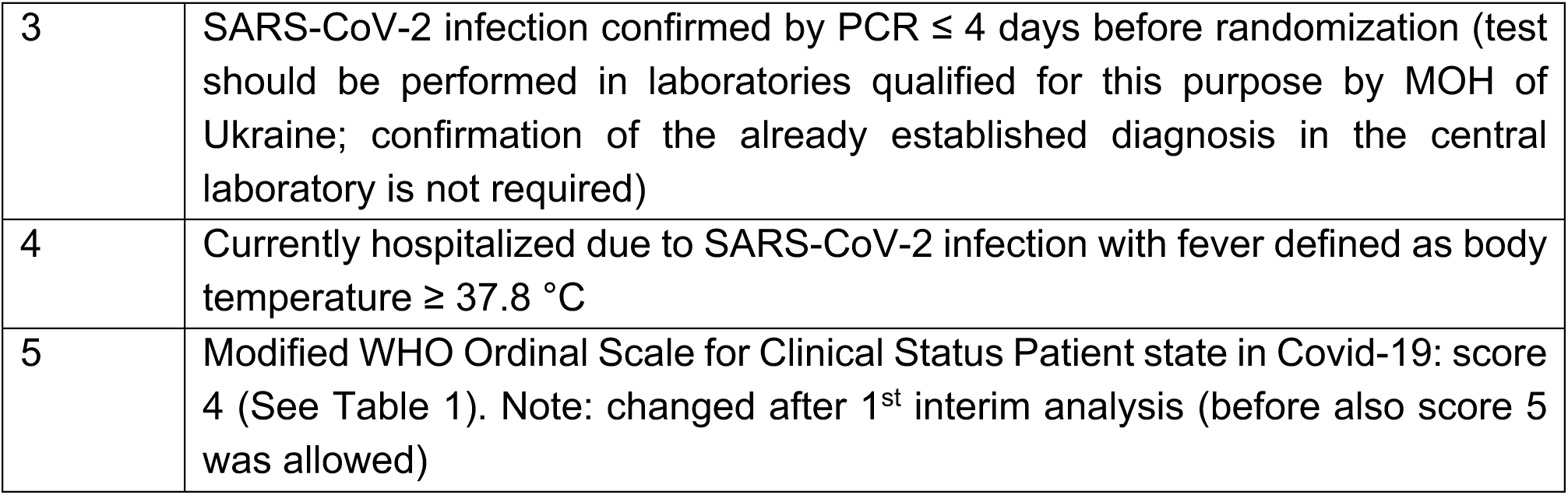
Patient inclusion criteria.

**Table 3:** Patient exclusion criteria.

| Number | Exclusion criterion |
| --- | --- |
| 1 | Concurrent treatment with other medicine with actual or possible direct-acting antiviral activity against SARS-CoV-2 is prohibited <24 hours prior to the start of enisamium or placebo treatment. |
| 2 | Requiring mechanical ventilation at screening or it is expected within 24 hours after inclusion. |
| 3 | Expected survival time <72 hours for any reason. |
| 4 | Positive pregnancy test. |
| 5 | Breastfeeding. |
| 6 | Presence of renal dysfunction defined as eGFR <60 mL/min, total bilirubin ≥2.0 mg/dL, TSH outside normal range and / or ASAT / ALAT above threefold upper limit of normal range (known from patient's medical history; after results from safety laboratory at visit 1 are available and this exclusion criterion would apply, this patient discontinues the clinical trial). |
| 7 | Known hypersensitivity to the trial drug, the metabolites, or formulation excipient. |
| 8 | History or presence of drug or alcohol abuse. |
| 9 | History or presence of diseases of thyroid gland. |
| 10 | Parallel participation in another clinical trial with an investigational product, participation in a clinical trial within less than 6 weeks prior to visit 1. |
| 11 | Known to be or suspected of being unable to comply with the trial protocol (e.g., no permanent address, history of drug abuse, known to be non-compliant or presenting an unstable psychiatric history). |
| 12 | Legal incapacity and / or other circumstances render the subject unable to understand the trial's nature, scope, and possible impact. |
| 13 | Subject in custody by juridical or official order. |
| 14 | Subject who has difficulties in understanding the language (Ukrainian) in which the subject information (informed consent form) is given. |
| 15 | Subjects who are members of the staff of the trial center, staff of the sponsor or the clinical research organization (CRO), the investigator him- / herself or close relatives of the investigator. |

### Randomization

At the screening stage, SARS-CoV-2 infection was confirmed by RT-qPCR in hospitalized patients with a diagnosis of COVID-19. Patients who met the criteria for inclusion and did not meet the criteria for exclusion were randomized 1:1 into a group receiving oral administration of enisamium and a group receiving oral administration of placebo. In addition, all patients received any other treatment deemed necessary by the investigator depending on the patient health status (“standard of care”). The first dose of enisamium or placebo was administered after the randomization procedure on visit 1 (day 1). Patients subsequently received enisamium or placebo 4 times a day (4x1 capsules) every 6 hours. The total treatment duration was 168 hours (7 or 8 days).

### Procedures

On day 1, patients were screened, randomized, and started on the treatment. On days 2 to 7 (or days 2 to 8, if the 168 hours of treatment ended on day 8), patients were treated, monitored, and evaluated, provided the patient had not been discharged from the hospital. Treatment with enisamium or placebo could be stopped if a patient was transferred to mechanical ventilation. On days 8 to 29, no treatment was given, but patients continued to be observed and evaluated for clinical symptoms if they had not yet been discharged from the hospital. For patients discharged before day 29, a follow-up visit was conducted on day 29 for final evaluation.

Throat swabs or sputum tests were taken for SARS-CoV-2 RNA detection by RT-qPCR on day 1, and if the patient remained hospitalized, on day 8 (+/- 1 day), day 15 (+/- 1 day), day 22 (+/- 1 day) and day 29 (+/- 1 day), or on the day of discharge from the hospital, if discharge occurred earlier than the planned RT-qPCR analysis points. During the study, routine throat swabs and sputum collections were performed for qualitative SARS-CoV-2 RNA detection by RT-qPCR according to the standard COVID-19 management protocols approved by the Ministry of Health of Ukraine. The samples were analyzed in laboratories accredited by the Ministry of Health of Ukraine. The RT-qPCR results were used to determine patient inclusion in the study, evaluate a patient’s clinical status during hospitalization, and decide the end point of hospitalization.

### Outcomes

The primary endpoint of this study was the time to improvement of the patient’s condition by 2 points (from 4 to 6) on the modified WHO scale (Table 1), as measured by assessment of the patient’s condition.

### Statistical analysis

All primary and safety analyses were based on the intention-to-treat (ITT) principle. The primary endpoint analysis was performed using Kaplan-Maier curves, Hazard function curves, medians of time to event, and logrank test. An analysis by patient age was not defined in the protocol when this was submitted early in the COVID-19 pandemic. However, as evidence for a correlation between age and COVID-19 severity accumulated over the course of the pandemic, we considered it prudent to consider age in our analysis and divided patients into age groups (“<40 years”, “40-<65 years” and “≥65 years”). For analysis of secondary endpoints descriptive statistics, graphics methods, exact Fisher test, logrank test, Cox regression and confidence intervals were used. For assessing the superiority of enisamium relative to placebo, we performed one-sided statistical hypothesis testing and set a significance level of 0.0131 for primary endpoints. A significance level of 0.025 was used to perform one-sided statistical hypotheses for secondary endpoints. Student’s t-tests were used to compare independent samples, while the Mann-Whitney test or Fisher’s exact test was used to test the initial homogeneity of the patient group, depending on the nature of the data and distribution. In the analysis of the initial homogeneity of the groups, a significance level of 0.05 (bilateral) was used. All statistical calculations were performed according to the principles of applied adaptive research design, and inflation of the level of significance was taken into account during the analysis.

### Interim analyses and protocol adjustments

To understand the effect of enisamium on the recovery of COVID-19 patients, a phase 3 clinical trial was started in 14 hospitals across the Ukraine in the summer of 2020. Since COVID-19 was insufficiently studied at the start of our study, it was difficult to predict the effect of enisamium on COVID-19 patients. We initially set out to study the effect of enisamium on patients with a baseline severity score (SR) of 4 or 5 on the modified WHO scale (Table 1) and randomized approximate 700 patients, to ensure we could include at least 398 patients in the ITT population (199 for each treatment group). In addition, we scheduled an conditional power (or interim) analysis based on the prospective approach of Mehta and Pocock, and Broberg to assess whether the ongoing study was sufficiently promising to reach the primary endpoint either the SR 4 or 5 group, or both, and whether more patients could be recruited for either group or whether the study should be terminated for one or both groups.^11, 12^ The conditional power analysis was conducted by an independent data monitoring committee.

When over 50% of the patients for the initial protocol design had been recruited (77 patients with a baseline of SR = 4 and 298 patients with a baseline of SR = 5 on the modified WHO scale), the preplanned conditional power analysis was performed. The interim analysis showed no promising result for the whole analysis population in the primary endpoint. Further subgroup investigations of interim data revealed that no relevant treatment difference could be observed in patients with a baseline of SR = 5. However, patients with a baseline of SR = 4, i.e., patients suffering from a higher degree of COVID-19 at baseline, showed a very promising benefit of enisamium treatment compared to placebo in the primary endpoint. Based on these interim results, recruitment of COVID-19 patients with a baseline of SR = 4 was continued to increase the sample size, whereas recruitment of COVID-19 patients who had a baseline of SR = 5 was stopped. This change was approved by the CEB and the Ethics Commissions at the treatment and prevention facilities where the study was conducted. We will refer to this latter group as the ITT population from here on.

In order to confirm that the observed trend would continue for the ITT group (and thereby protect the COVID-19 patients), a second interim analysis was planned after approximately 50% of the preplanned ITT population (i.e., 200 subjects with a baseline of SR = 4) had completed the study. This interim analysis was performed according to the two-step approach described by Bauer and Köhne.^13^ Following data clearance and adjustment of the statistical model based on the results of conditional power analysis, a one-sided p-value lower than the Bauer-Köhne cut-off of p = 0.0131 in the primary endpoint was needed to show a significant benefit of enisamium treatment. Since we found that p < 0.0131 between the enisamium and placebo populations in the second analysis, the clinical trial could be terminated.

Changes in clinical status are summarized on a continuous scale using means, standard deviations (SD), medians, interquartile ranges (IQR), and ranges. This trial was registered with ClinicalTrials.gov under NCT04682873.

## RESULTS

### Patients

A total of 592 patients were tested for the presence of SARS-CoV-2 RNA and randomized, of whom 296 were assigned to the placebo group and 296 to the enisamium group (Fig. 1). The safety and tolerability analysis (SA) population included all patients who received a dose of enisamium or placebo at least once, with 289 patients in the placebo group and 293 in the enisamium group, and thus a total of 582 patients. Given that after the first interim analysis, we decided to focus on subjects whose baseline score was SR = 4 and those who received at least one dose of enisamium in combination with standard care or placebo in combination with standard care (see Methods), the ITT population included 285 subjects, of which 143 subjects had been assigned to the placebo group and 142 subjects to the enisamium group. Some patients included in the ITT population met exclusion criterion 6 (Table 3) during the study and were subsequently excluded from further participation for safety reasons. These patients were included in the efficacy evaluation for the time they were in the study and received the study drug. The deviations from the original protocol as well as patient exclusions are illustrated in Fig. 1.

**Figure 1.**
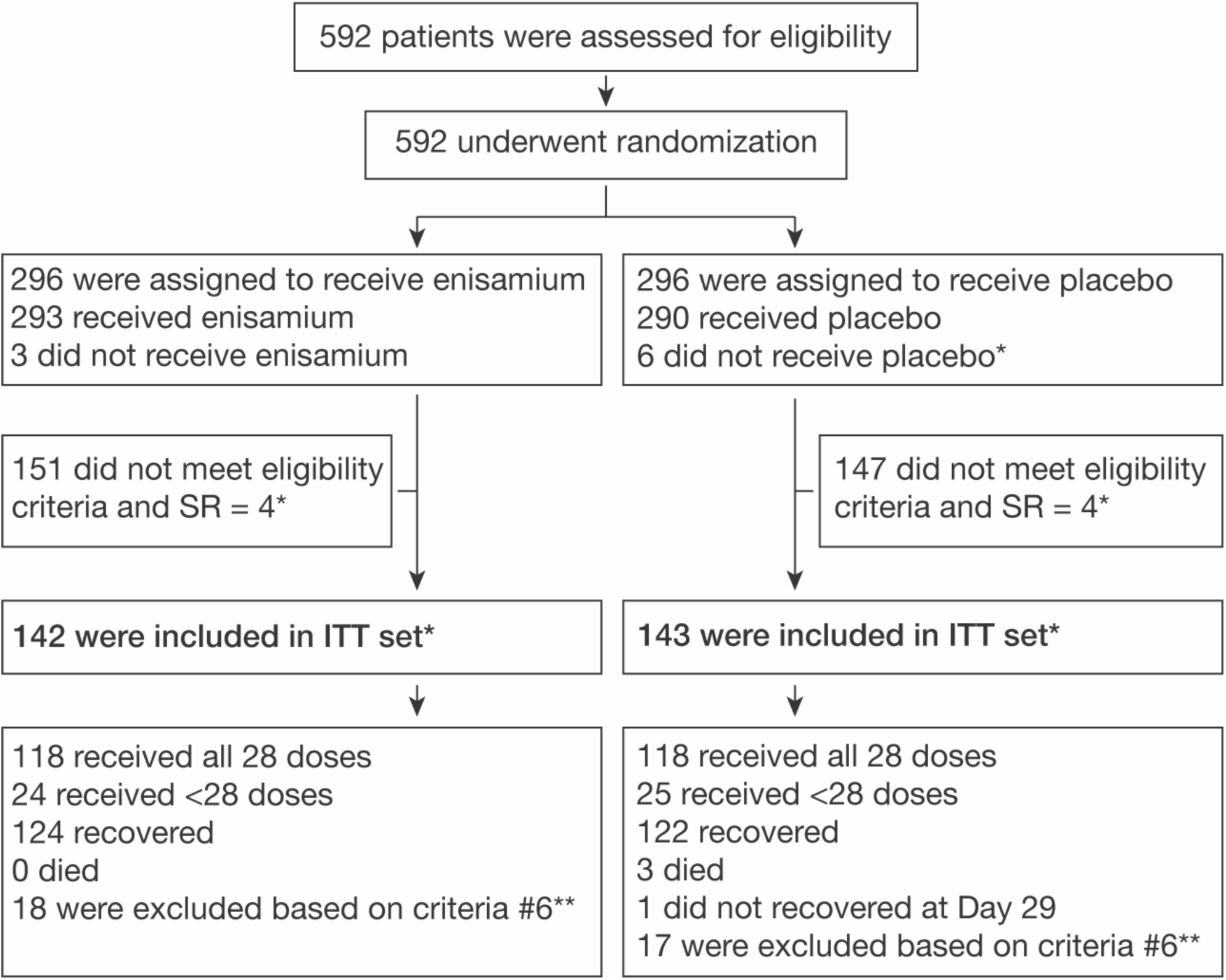
Schematic overview of patient recruitment, randomization, and treatment. *According to protocol amendment ITT set were defined as: “The ITT includes all randomized and IMP-treated subjects who had a Covid-19 subject severity rating (SR) of score 4 on Day 1 at time of randomization, and have valid post-baseline efficacy data”. **Presence of renal dysfunction defined as eGFR <60 mL/min, total bilirubin ≥2.0 mg/dL, TSH outside normal range and / or ASAT / ALAT above threefold upper limit of normal range (known from patient’s medical history; after results from safety laboratory at visit 1 are available (the results of the analysis could be known in a few days) and this exclusion criterion would apply, this patient discontinues the clinical trial.)

The ITT population was 47.0% male and 53.0% female (Table 4). The median age of the ITT population was 59 years (IQR 47–65), and across the three age groups 11.2 % were <40 years, 61,1 % were 40 –<65 years and 27.7 % were ≥65 years. All patients were from Ukraine, 99.6 % - Caucasian (white). Most patients had either one (36.5%), two or more (26.7%) of the prespecified coexisting conditions at enrollment. The most common comorbidities were hypertension (49.1%), BMI ≥30 kg/m^2^ (33.0%), and type 2 diabetes mellitus (9.1%). The median number of days between symptom onset and randomization was 8 days (IQR 6–12).

**Table 4.** Demographic and Clinical Characteristics at Baseline for ITT set*.

| Characteristic | All<br>N=285 | Enisamium<br>N=142 | Placebo<br>N=143 |
| --- | --- | --- | --- |
| Age, years — Median (IQR) | 59 (47 – 65) | 59 (47 – 65) | 59 (47.5 – 65) |
| Age category — no. (%) |  |  |  |
| — <40 year | 32 (11.2) | 15 (10.6) | 17 (11.9) |
| — 40 –<65 years | 174 (61.1) | 89 (62.7) | 85 (59.4) |
| — ≥65 years | 79 (27.7) | 38 (26.8) | 41 (28.7) |
| Sex — no. (%) |  |  |  |
| — male | 134 (47.0) | 64 (45.1) | 70 (49.0) |
| — female | 151 (53.0) | 78 (54.9) | 73 (51.0) |
| Race or ethnic group — no. (%)** |  |  |  |
| — Caucasian (white) | 284 (99.6) | 141 (99.3) | 143 (100.0) |
| — Asian | 1 (0.4) | 1 (0.7) | 0 (0.0) |
| Median time (IQR) from symptom onset to randomization, days | 8 (6 – 12) | 8 (5 – 10) | 7 (5 – 9) |
| No. of coexisting conditions — no. (%) |  |  |  |
| None | 105 (36.8) | 55 (38.7) | 50 (35.0) |
| One | 104 (36.5) | 52 (36.6) | 52 (36.4) |
| Two or more | 76 (26.7) | 35 (24.6) | 41 (28.7) |
| Coexisting conditions — no. (%) |  |  |  |
| Hypertension | 140 (49.1) | 67 (47.2) | 73 (51.5) |
| BMI $\geq 30$ kg/m<sup>2</sup> | 94 (33.0) | 47 (33.1) | 47 (32.9) |
| Type 2 diabetes | 26 (9.1) | 10 (7.0) | 16 (11.2) |
| * Percentages may not total to 100 because of rounding. |  |  |  |
| ** Race and ethnic group were reported by the patients. |  |  |  |

### Primary Endpoint

The ITT population consisted of 285 COVID-19 patients with a baseline SR of 4 points on the modified WHO scale (Table 1). The point of clinical improvement was set at an SR of 6 points. When estimating the number of days before reaching the moment of clinical improvement, all days were counted, including the 1^st^ day of the patient’s stay in the study. The day on which the patient’s condition reached an SR of 6 points was not included. Patients who died (1 point on the modified WHO scale) were considered in the analysis as those who did not achieve clinical improvement during the entire observation period (29 days). Similarly, patients who did not achieve clinical improvement (i.e., remained in a stable condition, improved in their condition by only 1 point, or declined in their condition) within 28 days were considered as patients who remained in the study for 29 days. Over the course of the study, the patients in the enisamium group all survived, while in the placebo group 3 patients died and 1 patient remained in SR = 4 on day 29 of the study.

Overall, patients in the enisamium group reached the primary endpoint after a median of 10 days compared to a median of 11 days for patients in the placebo group (Fig. 2 and Table 5). Because the differences between the enisamium and placebo group were significant (p<0.0131), the study was stopped in accordance with the protocol. These differences were also present and significant in the stratified age categories (one-sided logrank p = 0.00945). Patients <40 years reached the primary endpoint with a median of 8 days in the enisamium group and 9 days in the placebo group. In the 40-<65 years group, the medians were 10 and 11 days, respectively, and in the ≥65 years group 10 and 12 days, respectively.

**Figure 2.**
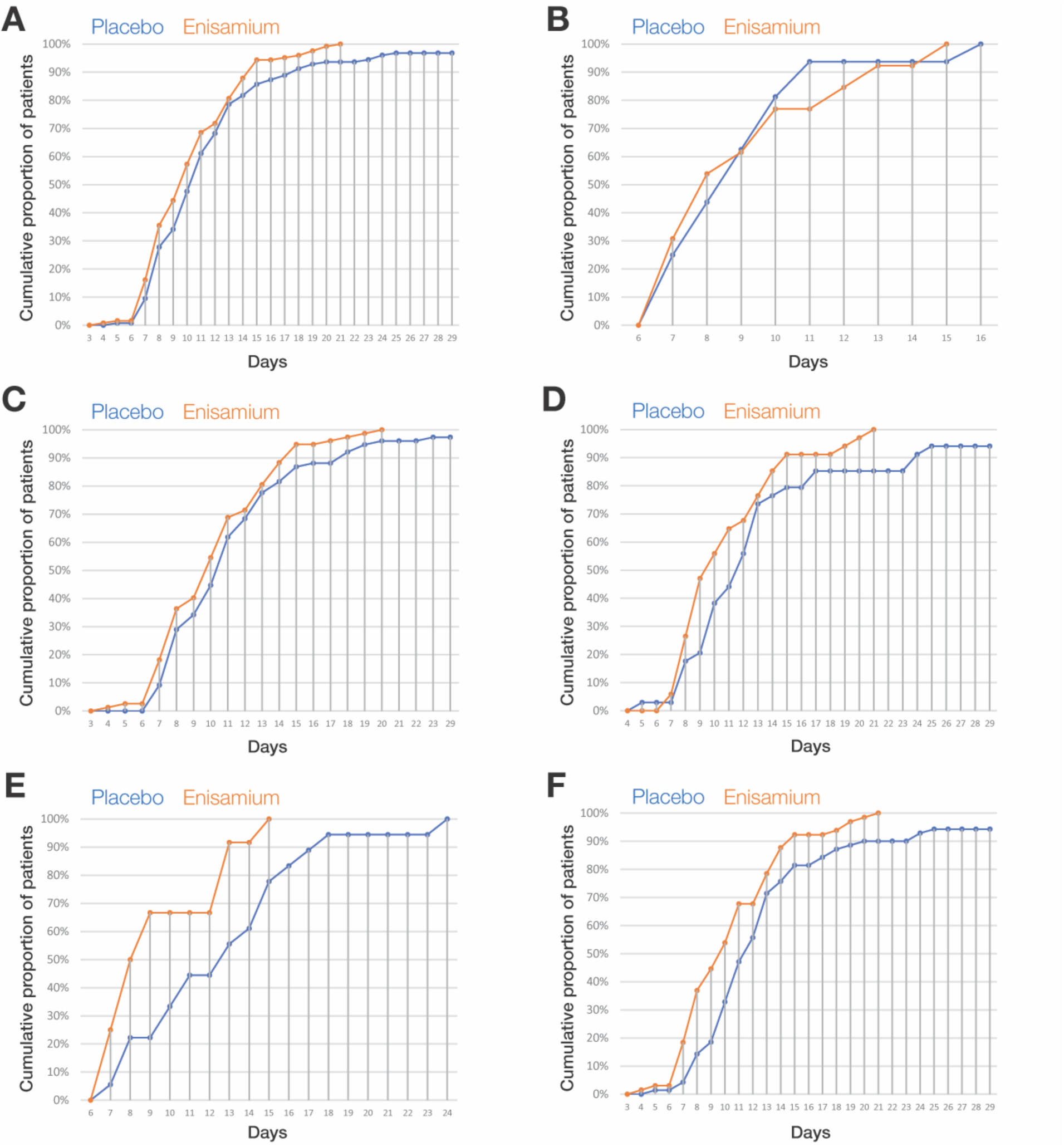
Kaplan-Meier plots. Kaplan-Meier estimates of the cumulative proportion of patients that achieved the primary endpoint in A) the overall patient population (n = 258), B) patients in the age category “< 40 years” (n = 32), C) patients in the age category “40 – < 65 years” (n = 174), D) patients in the age category “≥ 65 years” (n = 79), E) patients aged ≥ 50 years randomized within <5 days of symptom onset (n = 33), and F) patients aged >= 50 years randomized within <10 days of symptom onset (n = 154).

**Table 5.** Median time to reach primary endpoints for all patients and patient subgroups.

| Population |  | Group | n | Median |  |  |  | P-value<br>(one-sided) |
| --- | --- | --- | --- | --- | --- | --- | --- | --- |
|  |  |  |  | Estimate | Std.<br>Error | 95% CI |  |  |
|  |  |  |  |  |  | Lower<br>Bound | Upper<br>Bound |  |
| All ITT<br>patients (SR<br>= 4 at<br>baseline)<br>with age<br>stratification | Patients <40<br>years | Placebo | 17 | 9 | 0.65 | 7.73 | 10.27 | 0.009* |
|  |  | Enisamium | 15 | 8 | 0.90 | 6.24 | 9.76 |  |
|  |  | Total | 32 | 9 | 0.52 | 7.98 | 10.02 |  |
|  | Patients 40-<br><65 years | Placebo | 85 | 11 | 0.33 | 10.36 | 11.64 |  |
|  |  | Enisamium | 89 | 10 | 0.40 | 9.22 | 10.78 |  |
|  |  | Total | 174 | 11 | 0.27 | 10.46 | 11.54 |  |
|  | Patients ≥65<br>years | Placebo | 41 | 12 | 0.72 | 10.58 | 13.42 |  |
|  |  | Enisamium | 38 | 10 | 0.58 | 8.87 | 11.13 |  |
|  |  | Total | 79 | 11 | 0.65 | 9.73 | 12.27 |  |
| All patients randomized<br>within <5 days of symptom<br>onset |  | Placebo | 18 | 13 | 2.11 | 8.87 | 17.13 | 0.005 |
|  |  | Enisamium | 15 | 8 | 0.69 | 6.64 | 9.36 |  |
|  |  | Total | 33 | 10 | 1.10 | 7.85 | 12.15 |  |
| Patients aged ≥50 years<br>randomized within <10<br>days of symptom onset |  | Placebo | 81 | 12 | 0.52 | 10.98 | 13.02 | 0.002 |
|  |  | Enisamium | 73 | 10 | 0.54 | 8.95 | 11.05 |  |
|  |  | Total | 154 | 11 | 0.30 | 10.41 | 11.59 |  |

\* obtained using a logrank test that was stratified by age category

Among patients who were randomized <5 days after symptom onset, the enisamium group reached the primary endpoint after a median of 8 days compared to a median of 13 days for patients in the placebo group (p = 0.0051; 33 patients (Fig. 2 and Table 5). Among patients aged ≥50 years who had been randomized within <10 days of symptom onset, the enisamium group needed a median of 10 days to reach the primary endpoint whereas patients in the placebo group required a median of 12 days (one-sided p = 0.002; 154 patients) (Fig. 2 and Table 5).

### Secondary endpoints

During the trial, we recorded secondary endpoints including “discharge from hospital on day 15”, “discharge from hospital on day 22”, and “prevention of deterioration after randomization”. We observed that on day 15, significantly fewer patients remained in the hospital in the enisamium-treated group compared to the placebo group (5.65% vs. 14.29%; one-sided p = 0.018) (Table 6). On day 22, there were 0% hospitalized patients in the enisamium group compared to 6.3% the placebo group (0.0% vs. 6.3%; one-sided p = 0.004).

**Table 6.** Results for secondary endpoints by category.

| Variable | Category | Placebo |  | Amizon<sup>®</sup> MAX |  | P-value (one-sided) |
| --- | --- | --- | --- | --- | --- | --- |
|  |  | n | % | n | % |  |
| Patients discharged on day 8 | Patient not discharged | 92 | 73,0 | 81 | 65,3 | 0.119 |
| Patients discharged on day 15 | Patient not discharged | 18 | 14.3 | 7 | 5.6 | 0.018 |
| Patients discharged on day 22 | Patient not discharged | 8 | 6.3 | 0 | 0.0 | 0.004 |
| Patients discharged on day 29 | Patient not discharged | 4 | 3,2 | 0 | 0,0 | 0.063 |
| Deterioration by 1 SR point | Patients with deterioration of 1 point by SR scale | 12 | 8.4 | 3 | 2.1 | 0.016 |

| Variable | Category | Placebo |  | Amizon® MAX |  | P-value (one-sided) |
| --- | --- | --- | --- | --- | --- | --- |
|  |  | n | % | n | % |  |
| RT-qPCR test results on day 8 | SARS-CoV-2 positive on day 8 | 60 | 51.3 | 52 | 45.6 | 0.233 |
| RT-qPCR test results on day 15 | SARS-CoV-2 positive on day 15 | 8 | 9.2 | 5 | 5.6 | 0.289 |
| RT-qPCR test results on day 22 | SARS-CoV-2 positive on day 22 | 2 | 2.4 | 0 | 0.0 | 0.289 |
| RT-qPCR test results on day 29 | SARS-CoV-2 positive on day 29 | 1 | 1.2 | 0 | 0.0 | 0.494 |
| Mortality | Diseased patients | 3 | 2.1 | 0 | 0.0 | 0.125 |

**Table 7.** Results for the secondary endpoints by study group.

| Variable | Group | Statistics |  |  |  |  |  | P-value (one-sided) |
| --- | --- | --- | --- | --- | --- | --- | --- | --- |
|  |  | n | M | Me | SD | MIN | MAX |  |
| The sum of the scores of the subject's condition from the 2nd to the 15th day (SSR-15), points | Placebo | 143 | 70,80 | 72 | 11,98 | 22 | 90 | 0,079 |
|  | Amizon® MAX | 142 | 72,94 | 75,5 | 11,09 | 47 | 91 |  |
| The sum of the scores of the subject's condition from the 2nd to the 29th day (SSR-29), points | Placebo | 143 | 170,35 | 181 | 31,88 | 36 | 206 | 0,037 |
|  | Amizon® MAX | 142 | 175,85 | 186 | 26,71 | 112 | 203 |  |
| Assessment of the subject's condition on the 15th day (SR-15), points | Placebo | 143 | 6,64 | 7 | 1,651 | 1 | 8 | 0,137 |
|  | Amizon® MAX | 142 | 6,85 | 8 | 1,48 | 4 | 8 |  |
| Assessment of the condition of the subject on the 29th day (SR-29), points | Placebo | 143 | 7,27 | 8 | 1,589 | 1 | 8 | 0,109 |
|  | Amizon® MAX | 142 | 7,46 | 8 | 1,303 | 4 | 8 |  |
| Days alive and out of Hospital until Day 15 (DAOH-14), days | Placebo | 143 | 2,85 | 3 | 2,551 | 0 | 9 | 0,112 |
|  | Amizon® MAX | 142 | 3,25 | 3 | 2,740 | 0 | 10 |  |
| Estimate of global efficacy by investigator, scores | Placebo | 124 | 0,99 | 1 | 0,393 | 0 | 3 | 0,168 |
|  | Amizon® MAX | 123 | 0,93 | 1 | 0,248 | 0 | 1 |  |
| Estimate of global efficacy by subjects, scores | Placebo | 124 | 0,90 | 1 | 0,484 | 0 | 3 | 0,374 |
|  | Amizon® MAX | 123 | 0,86 | 1 | 0,347 | 0 | 1 |  |

**Table 8.**
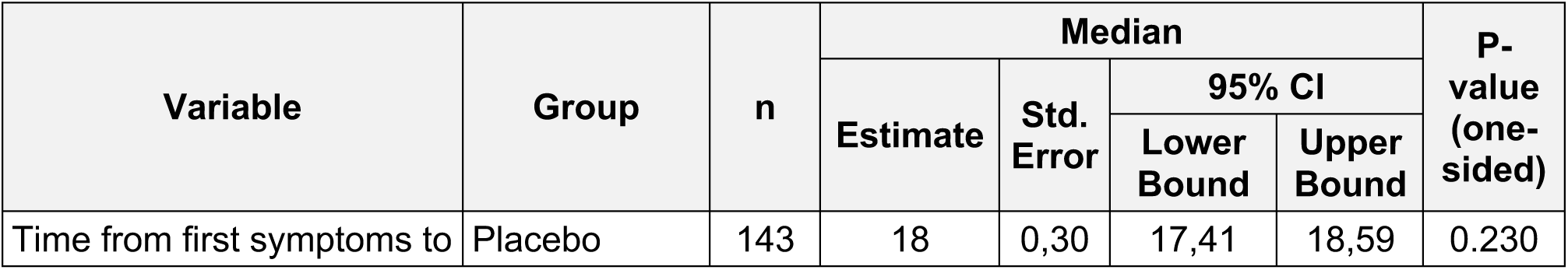
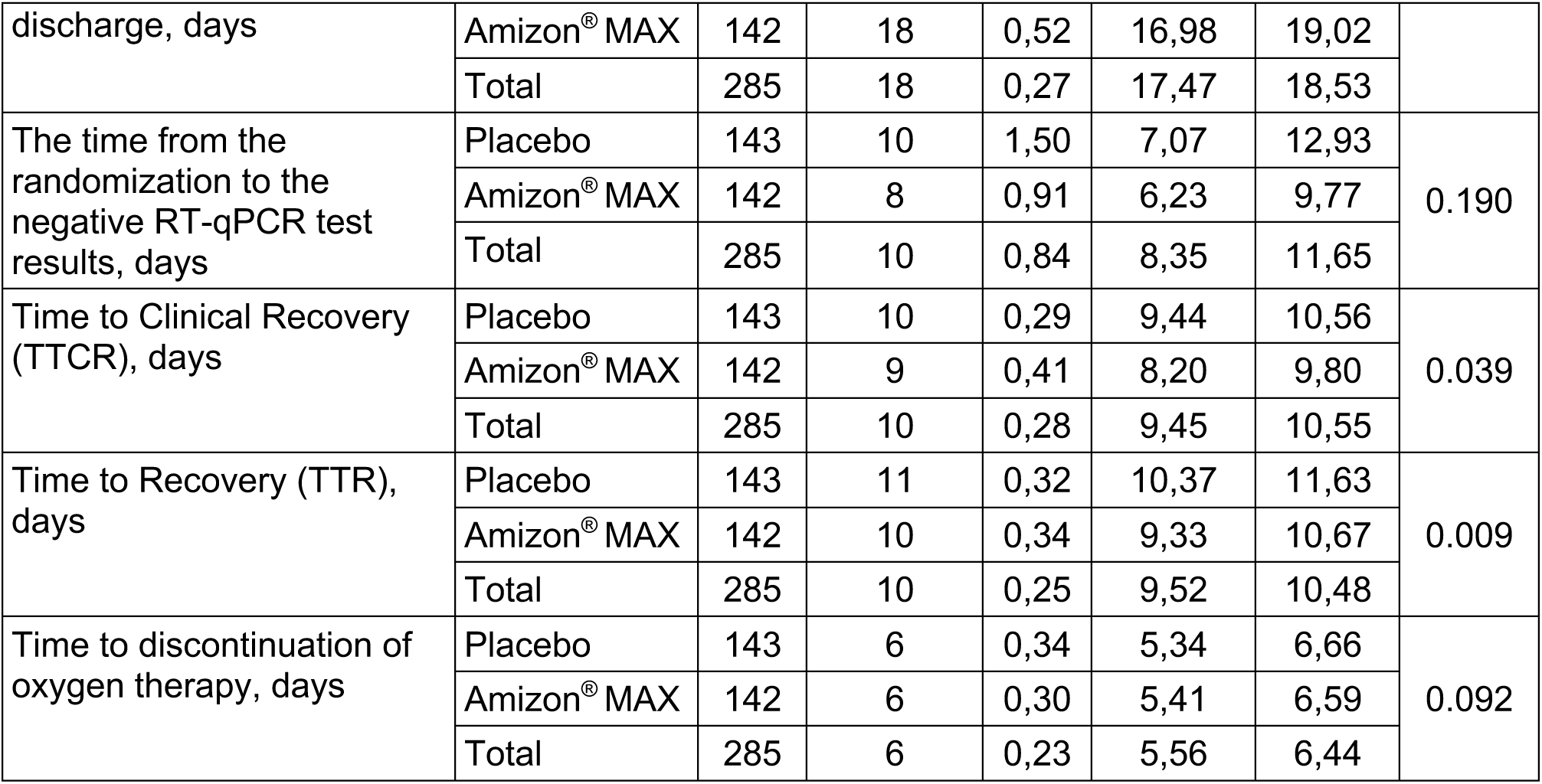
Results for secondary endpoints by time to event.

| Variable | Group | n | Median |  |  |  | P-value<br>(one-sided) |
| --- | --- | --- | --- | --- | --- | --- | --- |
|  |  |  | Estimate | Std.<br>Error | 95% CI |  |  |
|  |  |  |  |  | Lower<br>Bound | Upper<br>Bound |  |
| Time from first symptoms to | Placebo | 143 | 18 | 0,30 | 17,41 | 18,59 | 0.230 |
| discharge, days | Amizon® MAX | 142 | 18 | 0,52 | 16,98 | 19,02 |  |
|  | Total | 285 | 18 | 0,27 | 17,47 | 18,53 |  |
| The time from the randomization to the negative RT-qPCR test results, days | Placebo | 143 | 10 | 1,50 | 7,07 | 12,93 | 0.190 |
|  | Amizon® MAX | 142 | 8 | 0,91 | 6,23 | 9,77 |  |
|  | Total | 285 | 10 | 0,84 | 8,35 | 11,65 |  |
| Time to Clinical Recovery (TTCR), days | Placebo | 143 | 10 | 0,29 | 9,44 | 10,56 | 0.039 |
|  | Amizon® MAX | 142 | 9 | 0,41 | 8,20 | 9,80 |  |
|  | Total | 285 | 10 | 0,28 | 9,45 | 10,55 |  |
| Time to Recovery (TTR), days | Placebo | 143 | 11 | 0,32 | 10,37 | 11,63 | 0.009 |
|  | Amizon® MAX | 142 | 10 | 0,34 | 9,33 | 10,67 |  |
|  | Total | 285 | 10 | 0,25 | 9,52 | 10,48 |  |
| Time to discontinuation of oxygen therapy, days | Placebo | 143 | 6 | 0,34 | 5,34 | 6,66 | 0.092 |
|  | Amizon® MAX | 142 | 6 | 0,30 | 5,41 | 6,59 |  |
|  | Total | 285 | 6 | 0,23 | 5,56 | 6,44 |  |

We also kept track of the need for oxygen support. The need for stronger oxygen support was assessed by calculating the frequency and time to deterioration of the patient by 1 point on the modified WHO scale (i.e., from SR = 4 to SR = 3 points), as patients with the deteriorated condition had to receive non-invasive or high-flow oxygen therapy. The proportion of patients who deteriorated was 8.4% in the placebo group and 2.1% in the enisamium group, which is significant better in the enisamium group compared to the placebo group (p = 0.016; one-sided). If we evaluate the ratio of the chances of preventing the deterioration of the patient’s condition in the enisamium group compared with the placebo group, we obtain an – OR = 4.244 (95% CI: 1.171 – 15.380). Thus, we conclude that the use of enisamium increases the chances of preventing deterioration of patients by about 4 times compared with placebo. Finally, we observed fewer SARS-CoV-2 RT-qPCR results in the enisamium group compared to the placebo group at day 15, but this difference was not statistically significant (5.6 % vs. 9.2 %; one-sided p = 0.269). The results for abovementioned secondary endpoints end other secondary endpoints are listed in Tables 6 – 8.

### Symptom dynamics

At the start of the study, 98.4% of subjects in the placebo group and 97.2% of subjects in the enisamium group had a cough of varying severity (mild, moderate, or severe). On days 3, 4, and 5 after initiation of treatment, the proportion of patients who demonstrated decreased cough severity was statistically significantly higher in the enisamium group compared to the placebo group (day 3: 21.8% vs. 10.6%, p = 0.007; day 4: 33.8% vs. 21.1%, p = 0.011; day 5: 47.9% vs. 32.4%, p = 0.005). No statistically significant differences were observed on the other study days. In addition, we observed no statistically significant differences for other recorded symptoms, which included rhinorrhea, sore throat, headache, shortness of breath, diarrhea, myalgia, and fatigue (Table 9).

**Table 9.** Summary results for symptoms severity.

| Variable | Category | Placebo<br>(N = 142) |  | Amizon® MAX<br>(N = 142) |  | P-value<br>(one-sided) |
| --- | --- | --- | --- | --- | --- | --- |
|  |  | n | % | n | % |  |
| Cough severity on the day 2 | Reduced severity | 10 | 7,0 | 12 | 8,5 | 0,412 |
| Cough severity on the day 3 | Reduced severity | 15 | 10.5 | 31 | 21.8 | 0.008 |
| Cough severity on the day 4 | Reduced severity | 30 | 21.0 | 48 | 33.8 | 0.012 |
| Cough severity on the day 5 | Reduced severity | 46 | 32.2 | 68 | 47.8 | 0.005 |
| Cough severity on the day 6 | Reduced severity | 69 | 48,6 | 76 | 53,5 | 0.238 |
| Cough severity on the day 7 | Reduced severity | 78 | 54,9 | 85 | 59,9 | 0.236 |
| Cough severity on the day 8 | Reduced severity | 83 | 58,5 | 95 | 66,9 | 0.089 |
| Cough severity on the day 9 | Reduced severity | 89 | 62,7 | 97 | 68,3 | 0.191 |
| Cough severity on the day 10 | Reduced severity | 92 | 64,8 | 99 | 69,7 | 0.224 |
| Cough severity on the day 11 | Reduced severity | 95 | 66,9 | 98 | 69,0 | 0.400 |
|  |  | n | % | n | % |  |
| Cough severity on the day 12 | Reduced severity | 96 | 67,6 | 101 | 71,1 | 0.303 |
| Cough severity on the day 13 | Reduced severity | 97 | 68,3 | 105 | 73,9 | 0.180 |
| Cough severity on the day 14 | Reduced severity | 98 | 69,0 | 108 | 76,1 | 0.116 |
| Cough severity on the day 15 | Reduced severity | 101 | 71,1 | 108 | 76,1 | 0.210 |
| Shortness of breath severity on the day 2 | Reduced severity | 14 | 9,9 | 12 | 8,5 | 0,419 |
| Shortness of breath severity on the day 3 | Reduced severity | 36 | 25,4 | 34 | 23,9 | 0,445 |
| Shortness of breath severity on the day 4 | Reduced severity | 69 | 48,6 | 62 | 43,7 | 0,238 |
| Shortness of breath severity on the day 5 | Reduced severity | 82 | 57,7 | 86 | 60,6 | 0,359 |
| Shortness of breath severity on the day 6 | Reduced severity | 95 | 66,9 | 97 | 68,3 | 0,450 |
| Shortness of breath severity on the day 7 | Reduced severity | 98 | 69,0 | 104 | 73,2 | 0,256 |
| Shortness of breath severity on the day 8 | Reduced severity | 104 | 73,2 | 113 | 79,6 | 0,132 |
| Shortness of breath severity on the day 9 | Reduced severity | 107 | 75,4 | 114 | 80,3 | 0,196 |
| Shortness of breath severity on the day 10 | Reduced severity | 111 | 78,2 | 115 | 81,0 | 0,330 |
| Shortness of breath severity on the day 11 | Reduced severity | 114 | 80,3 | 117 | 82,4 | 0,380 |
| Shortness of breath severity on the day 12 | Reduced severity | 113 | 79,6 | 121 | 85,2 | 0,138 |
| Shortness of breath severity on the day 13 | Reduced severity | 117 | 82,4 | 125 | 88,0 | 0,121 |
| Shortness of breath severity on the day 14 | Reduced severity | 117 | 82,4 | 126 | 88,7 | 0,088 |
| Shortness of breath severity on the day 15 | Reduced severity | 117 | 82,4 | 127 | 89,4 | 0,062 |
| Fatigue severity on the day 2 | Reduced severity | 17 | 12,0 | 13 | 9,2 | 0,282 |
| Fatigue severity on the day 3 | Reduced severity | 29 | 20,4 | 37 | 26,1 | 0,163 |
| Fatigue severity on the day 4 | Reduced severity | 51 | 35,9 | 58 | 40,8 | 0,232 |
| Fatigue severity on the day 5 | Reduced severity | 64 | 45,1 | 74 | 52,1 | 0,143 |
| Fatigue severity on the day 6 | Reduced severity | 84 | 59,2 | 85 | 59,9 | 0,500 |
| Fatigue severity on the day 7 | Reduced severity | 94 | 66,2 | 96 | 67,6 | 0,450 |
| Fatigue severity on the day 8 | Reduced severity | 101 | 71,1 | 105 | 73,9 | 0,345 |
| Fatigue severity on the day 9 | Reduced severity | 107 | 75,4 | 109 | 76,8 | 0,445 |
| Fatigue severity on the day 10 | Reduced severity | 114 | 80,3 | 109 | 76,8 | 0,282 |
| Fatigue severity on the day 11 | Reduced severity | 117 | 82,4 | 110 | 77,5 | 0,277 |
| Fatigue severity on the day 12 | Reduced severity | 117 | 82,4 | 114 | 80,3 | 0,380 |
|  |  | n | % | n | % |  |
| Fatigue severity on the day 13 | Reduced severity | 120 | 84,5 | 117 | 82,4 | 0,375 |
| Fatigue severity on the day 14 | Reduced severity | 122 | 85,9 | 119 | 83,8 | 0,370 |
| Fatigue severity on the day 15 | Reduced severity | 122 | 85,9 | 120 | 84,5 | 0,434 |
| Rhinorrhea severity on the day 2 | Reduced severity | 8 | 5,6 | 5 | 3,5 | 0,286 |
| Rhinorrhea severity on the day 3 | Reduced severity | 13 | 9,2 | 10 | 7,0 | 0,332 |
| Rhinorrhea severity on the day 4 | Reduced severity | 15 | 10,6 | 13 | 9,2 | 0,421 |
| Rhinorrhea severity on the day 5 | Reduced severity | 17 | 12,0 | 13 | 9,2 | 0,282 |
| Rhinorrhea severity on the day 6 | Reduced severity | 18 | 12,7 | 12 | 8,5 | 0,167 |
| Rhinorrhea severity on the day 7 | Reduced severity | 18 | 12,7 | 14 | 9,9 | 0,287 |
| Rhinorrhea severity on the day 8 | Reduced severity | 18 | 12,7 | 14 | 9,9 | 0,287 |
| Rhinorrhea severity on the day 9 | Reduced severity | 19 | 13,4 | 14 | 9,9 | 0,230 |
| Rhinorrhea severity on the day 10 | Reduced severity | 19 | 13,4 | 14 | 9,9 | 0,230 |
| Rhinorrhea severity on the day 11 | Reduced severity | 19 | 13,4 | 14 | 9,9 | 0,230 |
| Rhinorrhea severity on the day 12 | Reduced severity | 19 | 13,4 | 14 | 9,9 | 0,230 |
| Rhinorrhea severity on the day 13 | Reduced severity | 19 | 13,4 | 14 | 9,9 | 0,230 |
| Rhinorrhea severity on the day 14 | Reduced severity | 19 | 13,4 | 14 | 9,9 | 0,230 |
| Rhinorrhea severity on the day 15 | Reduced severity | 19 | 13,4 | 14 | 9,9 | 0,230 |
| Headache severity on the day 2 | Reduced severity | 21 | 14,8 | 31 | 21,8 | 0,083 |
| Headache severity on the day 3 | Reduced severity | 38 | 26,8 | 50 | 35,2 | 0,079 |
| Headache severity on the day 4 | Reduced severity | 57 | 40,1 | 65 | 45,8 | 0,201 |
| Headache severity on the day 5 | Reduced severity | 63 | 44,4 | 69 | 48,6 | 0,276 |
| Headache severity on the day 6 | Reduced severity | 69 | 48,6 | 72 | 50,7 | 0,406 |
| Headache severity on the day 7 | Reduced severity | 68 | 47,9 | 75 | 52,8 | 0,238 |
| Headache severity on the day 8 | Reduced severity | 75 | 52,8 | 80 | 56,3 | 0,317 |
|  |  | n | % | n | % |  |
| Headache severity on the day 9 | Reduced severity | 76 | 53,5 | 80 | 56,3 | 0,360 |
| Headache severity on the day 10 | Reduced severity | 73 | 51,4 | 83 | 58,5 | 0,142 |
| Headache severity on the day 11 | Reduced severity | 78 | 54,9 | 83 | 58,5 | 0,316 |
| Headache severity on the day 12 | Reduced severity | 78 | 54,9 | 84 | 59,2 | 0,275 |
| Headache severity on the day 13 | Reduced severity | 80 | 56,3 | 87 | 61,3 | 0,235 |
| Headache severity on the day 14 | Reduced severity | 80 | 56,3 | 89 | 62,7 | 0,167 |
| Headache severity on the day 15 | Reduced severity | 80 | 56,3 | 89 | 62,7 | 0,167 |
| Sore throat severity on the day 2 | Reduced severity | 15 | 10,6 | 12 | 8,5 | 0,343 |
| Sore throat severity on the day 3 | Reduced severity | 31 | 21,8 | 27 | 19,0 | 0,330 |
| Sore throat severity on the day 4 | Reduced severity | 42 | 29,6 | 35 | 24,6 | 0,212 |
| Sore throat severity on the day 5 | Reduced severity | 42 | 29,6 | 42 | 29,6 | 0,552 |
| Sore throat severity on the day 6 | Reduced severity | 44 | 31,0 | 45 | 31,7 | 0,500 |
| Sore throat severity on the day 7 | Reduced severity | 49 | 34,5 | 49 | 34,5 | 0,550 |
| Sore throat severity on the day 8 | Reduced severity | 52 | 36,6 | 49 | 34,5 | 0,402 |
| Sore throat severity on the day 9 | Reduced severity | 52 | 36,6 | 50 | 35,2 | 0,451 |
| Sore throat severity on the day 10 | Reduced severity | 54 | 38,0 | 53 | 37,3 | 0,500 |
| Sore throat severity on the day 11 | Reduced severity | 55 | 38,7 | 53 | 37,3 | 0,451 |
| Sore throat severity on the day 12 | Reduced severity | 55 | 38,7 | 55 | 38,7 | 0,548 |
| Sore throat severity on the day 13 | Reduced severity | 55 | 38,7 | 56 | 39,4 | 0,500 |
| Sore throat severity on the day 14 | Reduced severity | 55 | 38,7 | 57 | 40,1 | 0,452 |
| Sore throat severity on the day 15 | Reduced severity | 56 | 39,4 | 57 | 40,1 | 0,500 |
| Diarrhoea severity on the day 2 | Reduced severity | 10 | 7,0 | 8 | 5,6 | 0,404 |
| Diarrhoea severity on the day | Reduced severity | 15 | 10,6 | 15 | 10,6 | 0,578 |
|  |  | n | % | n | % |  |
| 3 |  |  |  |  |  |  |
| Diarrhoea severity on the day 4 | Reduced severity | 16 | 11,3 | 16 | 11,3 | 0,574 |
| Diarrhoea severity on the day 5 | Reduced severity | 18 | 12,7 | 13 | 9,2 | 0,224 |
| Diarrhoea severity on the day 6 | Reduced severity | 20 | 14,1 | 18 | 12,7 | 0,431 |
| Diarrhoea severity on the day 7 | Reduced severity | 19 | 13,4 | 17 | 12,0 | 0,429 |
| Diarrhoea severity on the day 8 | Reduced severity | 19 | 13,4 | 17 | 12,0 | 0,429 |
| Diarrhoea severity on the day 9 | Reduced severity | 19 | 13,4 | 20 | 14,1 | 0,500 |
| Diarrhoea severity on the day 10 | Reduced severity | 19 | 13,4 | 19 | 13,4 | 0,569 |
| Diarrhoea severity on the day 11 | Reduced severity | 19 | 13,4 | 19 | 13,4 | 0,569 |
| Diarrhoea severity on the day 12 | Reduced severity | 20 | 14,1 | 19 | 13,4 | 0,500 |
| Diarrhoea severity on the day 13 | Reduced severity | 21 | 14,8 | 19 | 13,4 | 0,432 |
| Diarrhoea severity on the day 14 | Reduced severity | 22 | 15,5 | 19 | 13,4 | 0,368 |
| Diarrhoea severity on the day 15 | Reduced severity | 22 | 15,5 | 19 | 13,4 | 0,368 |
| Myalgia severity on the day 2 | Reduced severity | 29 | 20,4 | 25 | 17,6 | 0,325 |
| Myalgia severity on the day 3 | Reduced severity | 52 | 36,6 | 51 | 35,9 | 0,500 |
| Myalgia severity on the day 4 | Reduced severity | 80 | 56,3 | 69 | 48,6 | 0,117 |
| Myalgia severity on the day 5 | Reduced severity | 85 | 59,9 | 82 | 57,7 | 0,405 |
| Myalgia severity on the day 6 | Reduced severity | 92 | 64,8 | 85 | 59,9 | 0,231 |
| Myalgia severity on the day 7 | Reduced severity | 95 | 66,9 | 90 | 63,4 | 0,309 |
| Myalgia severity on the day 8 | Reduced severity | 96 | 67,6 | 92 | 64,8 | 0,353 |
| Myalgia severity on the day 9 | Reduced severity | 97 | 68,3 | 93 | 65,5 | 0,353 |
| Myalgia severity on the day 10 | Reduced severity | 96 | 67,6 | 94 | 66,2 | 0,450 |
| Myalgia severity on the day 11 | Reduced severity | 98 | 69,0 | 97 | 68,3 | 0,500 |
| Myalgia severity on the day 12 | Reduced severity | 97 | 68,3 | 97 | 68,3 | 0,551 |
| Myalgia severity on the day 13 | Reduced severity | 97 | 68,3 | 98 | 69,0 | 0,500 |
| Myalgia severity on the day 14 | Reduced severity | 98 | 69,0 | 98 | 69,0 | 0,551 |
| Myalgia severity on the day 15 | Reduced severity | 98 | 69,0 | 98 | 69,0 | 0,551 |

### Safety Outcomes

The SA population (289 patients in the placebo group and 293 in the enisamium group) included 582 randomized patients who had each received at least one dose of the study drug or placebo control. A total of 28 doses were taken by 80.6% of the patients in the placebo group and 80.2% of the patients in the enisamium group. The study found 172 adverse reactions/adverse events (AR/AE) in 87 placebo subjects and 229 AR/AE in 105 patients in the enisamium group (Table 10). Most AR/AE were mild and moderate in both the placebo and enisamium groups. The physician classified causation as “related” for 24.4% of AR/AE in the placebo group and for 48.9% of AR/AE in the enisamium group. The AR/AE in the enisamium group that were associated with the study drug were mild or moderate and did not require additional treatment, and therefore, the safety of enisamium can be considered good. The investigators rated the overall tolerability of enisamium as very good (45.3%), good (49.1%), and moderate (5.6%) in the randomized patients. Differences in tolerability estimates between the placebo and enisamium groups were not statistically significant (p = 0.289; two-sided). Patients rated the overall tolerability of enisamium as very good (50.6%), good (43.4%), and moderate (5.6%). Differences in tolerability estimates between the two groups were not statistically significant (p = 0.260; two-sided).

**Table 10.** Summary results for the safety and tolerability endpoints.

| Parameter | Placebo group<br>(N=289) n (%) | Enisamium<br>(N=293) n (%) |
| --- | --- | --- |
| Subjects evaluated for AR/AE analysis | 289 | 293 |
| Number of AR/AE | 172 | 229 |
| Patients with AR/AE | 87 (30.1) | 105 (35.8) |
| Number of SAE | 5 (2.9) | 4 (1.7) |
| Patients with SAE | 3 (1.04) | 4 (1.37) |
| Patients are excluded due to AR/AE | 15 (5.2) | 15 (5.1) |

N = the number of patients in the total analyzed; n = the number of patients who had events; In each line, patients were included only once.

The study did not reveal negative clinically significant dynamics of laboratory parameters of the complete blood count (CBC) and other blood parameters (leukocytes, erythrocytes, hemoglobin, hematocrit, lymphocytes, monocytes, neutrophils, eosinophils, basophils, platelets, mean corpuscal hemoglobin (MCH), mean corpuscal volume (MCV) and mean corpuscal hemoglobin concentration (MCHC)) in both groups. In addition, the study did not reveal clinically significant dynamics of laboratory parameters of the general analysis of urine and did not reveal clinically significant changes in laboratory parameters of biochemical blood tests (ALT, AST, glucose, total bilirubin, creatinine, cholesterol, LDL, GGT, potassium, sodium, calcium, triglycerides, free thyroxine, free thyroxine, free tridecin- reactive protein) in both groups. Changes in these indicators during the study were random, and statistically and clinically insignificant. Normalization of the laboratory parameters was also observed and attributed to improvement in the clinical condition.

## DISCUSSION

The aim of this study was to evaluate the clinical efficacy and safety of enisamium compared to placebo when administered orally in combination with standard care in hospitalized patients with moderate COVID-19 disease. No other studies have assessed the clinical efficacy of enisamium for treatment of COVID-19 so far. We previously investigated the effect of enisamium on influenza patients and found that enisamium treatment improved patient recovery when compared to a placebo control.^8^ Here, we observed that enisamium significantly improves the recovery of patients with a modified WHO baseline score of SR = 4, i.e. hospitalized patients receiving oxygen support, relative to patients receiving placebo. While we observed an effect that was significant for this specific group, further research is needed to confirm that treatment of COVID-19 patients will result in a clinical impact. Other treatments, including molnupiravir and paxlovid, are available in several countries. Enisamium has received approval for clinical use in Ukraine.

During our study, all patients had access to standard of care. No fatalities were observed among the enisamium group in the ITT population and all patients in the enisamium group reached the primary endpoint within 21 days. By contrast, 3 fatalities were recorded in the placebo group and some patients in the placebo group took longer to reach the primary endpoint. The best patient improvement to the primary endpoint was observed when enisamium was administered within 5 days, which is in line with observations for other antivirals, such remdesivir, which are also most efficacious when given early in infection.

Our study was double-blinded and conducted at 14 centers, limiting bias in the observed outcomes. In addition, care was taken to confirm SARS-CoV-2 infection in hospitalized patients using RT-qPCR before randomization. This ensured that our study tested the effect of enisamium on the clinical aspects of COVID-19 and that it was not limited by deciding patient enrollment solely on clinical diagnosis. However, we did not test for the presence of other respiratory viruses and microbes, and cannot rule out that some patients may have had secondary infections. Secondary infections have been rare among COVID- 19 patients, and we do not expect these to have impacted the described observations. The median age of our ITT population was 59, which is relatively young, but a fair reflection of COVID-19 patients with moderate disease. Severe COVID-19 is typically observed in senior people and the reported age distribution is, therefore, not a limitation of our study. In addition, we observed that enisamium treatment is safe to use in COVID-19 patients with moderate disease and that an orally administered treatment in capsules of 0.5 g four times a day for 8 days is well tolerated.

In our interim analysis, no significant effect of enisamium treatment was observed among patients with a modified SR of 5 (hospitalized but no additional oxygen support), suggesting that the effect of enisamium may be linked to a specific group of hospitalized patients who needs non-invasive oxygen support. Due to the rapidly changing medical landscape in 2020-2021 and best protect patient health, we stopped recruitment of patients with a modified SR of 5, and focused on recruitment of patients with a modified SR of 4. Subsequent analyses and statistical calculations were performed according to the principles of applied adaptive research design, and inflation of the level of significance was performed to correct for interim analysis. Extended

In summary, our data suggest that for COVID-19 patients that do not require supplementary oxygen (SR = 5), standard care is sufficient to aid recovery and enisamium does not offer significant clinical benefits. However, standard care in combination with enisamium treatment is significantly more effective than standard care in combination with placebo treatment in patients with moderate COVID-19 requiring additional oxygen (SR = 4), suggesting that enisamium treatment is a safe and useful addition to the current antivirals.

## ACKNOWLEDGMENTS

The authors thank Stefano Elli, Federico Sala, Cesare Costentino, and Marco Guerrini for the docking and molecular dynamics simulations and for contributions to structural analysis and in silico analyses reported elsewhere, which were invaluable for the design of this study. In addition, the authors would like to thank Alexander Walker, Haitian Fan, Jeremy Keown, Denisa Bojkova, Marco Bechtel, Xinjian Peng, Jindrich Cinatl, Jonathan Grimes and Ervin Fodor for contributions to in vitro experiments reported elsewhere, which were invaluable for making this study possible. The authors would also like to thank Bill Johnson, Miguel Muzzio, David Boltz for valuable results and discussions regarding toxicology studies performed at their facllity, and for Dr. Juergen Richt for helpful discussions regarding the antiviral efficacy of the test compound.

## Author contributions

O.H., L.M., V.M., and A.G. designed study. O.H., A.M., Y.L, L.M. and V.M. coordinated study. H.S., J.M., P.B., A.T.V performed data analysis and data presentation. P.B., A.T.V, and V.M. wrote manuscript. All authors read and approved the manuscript.

## Data availability

The data analyzed and presented in this study are available from P.B. and V.M. on reasonable request, providing the request meets local ethical and research criteria. Patient data will be anonymized, and study documents will be redacted to protect the privacy of trial participants. The study protocol is available upon request. The trial was registered with ClinicalTrials.gov under NCT04682873.

